# The Stability and Predictive Value of Negative Symptom Dimensions in First-Episode Psychosis: A 5-Year Follow-Up Study

**DOI:** 10.64898/2026.03.18.26348724

**Authors:** Yifei Lang, Tabea Schoeler, Giada Tripoli, Giulia Trotta, Victoria Rodriguez, Edoardo Spinazzola, Luis Alameda, Xinlin Li, Sagnik Bhattacharyya, Craig Morgan, Charlotte Gayer-Anderson, Valeria Mondelli, Simona A. Stilo, Antonella Trotta, Lucia Sideli, Olesya Ajnakina, Paola Dazzan, Fiona Gaughran, Anthony S. David, Marta Di Forti, Robin M. Murray, Diego Quattrone

**Author notes:** Correspondence concerning this article should be addressed to Yifei Lang,. Department of Psychosis Studies, Institute of Psychiatry, Psychology and Neuroscience (IoPPN), King’s College London, De Crespigny Park, Denmark Hill, London SE5 8AF, UK.

## Abstract

**Background:** Diminished Expression (DE) and Amotivation/Apathy (AA) are widely recognized as two main factors of negative symptoms. This study aimed to 1) examine the longitudinal stability of the DE-AA structure and its variation throughout a 5-year follow-up in people with first-episode psychosis (FEP), and 2) investigate whether DE and AA have distinct predictive value compared with the unitary construct of negative symptoms.

**Study Design:** 227 participants from the EUropean Network of National Schizophrenia Networks Studying Gene-Environment Interactions (EU-GEI) and Genetics and Psychosis (GAP) studies were included at FEP and were followed up 5 years later. One-factor (global negative symptoms), uncorrelated two-factor (DE-AA), and correlated two-factor structures were modelled using confirmatory factor analysis. Regression analyses were applied to examine the associations between these factors and negative symptom trajectories, functioning, and quality-of-life outcomes.

**Study Results:** The correlated two-factor model composed of DE and AA best fitted the data and exhibited 5-year stability. The regression model adjusted for AA accounted for more variance (59.2%) than global negative symptoms (52.8%) in explaining the enduring course of negative symptoms. Baseline AA was the only negative symptom factor that significantly predicted individuals’ functional outcome at follow-up (B=-1.76, p=0.037). All negative symptom dimensions negatively predicted employment status, whereas lower educational attainment was primarily related to AA severity at baseline.

**Conclusions:** Our findings support the validity and longitudinal stability of the two-dimensional (DE-AA) approach to negative symptoms in individuals with FEP. AA in particular exhibited distinctive predictive value, underscoring its potential clinical utility for early identification and the development of targeted interventions.

## Introduction

Given the strong associations with poor psychosocial functioning, reduced quality of life, and low remission rates (Hunter and Barry, 2012; Novick et al., 2009), negative symptoms are considered among the most disabling features of psychotic disorders, especially schizophrenia. Despite decades of research emphasizing the clinical importance of negative symptoms, effective treatments remain elusive (Fusar-Poli et al., 2015). One of the major challenges in addressing negative symptoms lies in their heterogeneity. Recent studies have highlighted that negative symptoms should not be viewed as a singular construct, but rather examined according to specific underlying dimensions (Galderisi et al., 2021).

Specifically, factor-analytic studies have consistently identified two negative symptom dimensions: Diminished Expression (DE) and Amotivation/Apathy (AA) (Strauss et al., 2013). These two factors have demonstrated robust measurement invariance in both first- and second-generation scales of negative symptoms (Jang et al., 2016; Li et al., 2022), and have shown distinct associations with clinical presentations and functional outcomes. For example, the AA dimension correlates with more severe positive symptoms, longer durations of hospitalization, and worse social functioning compared to DE (Harvey et al., 2017); while the DE subgroup has been linked to more pronounced cognitive impairments and greater difficulty with household activities (Galderisi et al., 2013; Hartmann-Riemer et al., 2015). According to Kirkpatrick (2014), these clusters of negative symptoms could be associated with different risk factors, illness progression, and perhaps should be considered separate treatment targets. Therefore, distinguishing between DE and AA in both clinical and research contexts may facilitate the development of targeted interventions and consequently improve the therapeutic outcome.

Given that the prevalence of at least one or more negative symptoms is notably high (30-70%) among individuals with first-episode psychosis (FEP) (Bobes et al., 2009; Downs et al., 2019; Malla et al., 2002), investigating their prognostic value in this early-stage population may yield clinical benefits. For example, it allows for timely interventions during the critical treatment window—the first year following the onset of psychosis—which has been proposed to be crucial in shaping the course of the disorder (Ihler et al., 2023). To date, however, the predictive value of negative symptom dimensions has primarily been explored in chronic samples (Galderisi et al., 2013; Peralta et al., 2014). The long-term equivalence of the DE-AA structure, including their associations with outcome variables, have yet to be thoroughly examined in FEP cohorts, even though negative symptoms may originate before or after the onset of the first episode (Peralta et al., 2000).

Furthermore, as enduring negative symptoms represent an unmet therapeutic challenge in schizophrenia, longitudinal studies examining symptom trajectories could be informative for prognosis prediction. For instance, the associations between baseline characteristics and enduring trajectory of the symptoms could reveal early indicators for the persistence of the disorder, which is typically associated with worse recovery outcomes (Chang et al., 2011). Therefore, early identification of individuals prone to developing enduring negative symptoms could be particularly beneficial to their rehabilitation efforts.

To summarize, this study aimed to 1) explore patterns of negative symptom change between baseline and 5-year follow-up in FEP, and their distinctions across sociodemographic, functional, and clinical variables at baseline and follow-up; 2) evaluate the factor structure of negative symptoms and their longitudinal equivalence; 3) investigate whether the DE-AA structure offers superior value compared to the global construct of negative symptoms in predicting functioning, hospitalization, and quality-of-life outcomes at the 5-year follow-up. In line with the decoupling paradigms of negative symptoms (Fernandez-Egea et al., 2023), we hypothesized that the DE-AA structure would remain stable over time, and their ability to predict outcome would differ and even surpass that of the global construct of negative symptoms in aforementioned respects.

## Methods

### Design and Participants

The dataset was drawn from the London branch of the EUropean network of national schizophrenia networks Gene-Environment Interactions (EU-GEI) study (Gayer-Anderson et al., 2020) and the Genetics and Psychosis (GAP) study (Di Forti et al., 2014; Murray et al., 2020), including patients from the South London & Maudsley Mental Health NHS Foundation Trust. Given that diagnoses of psychotic disorders may be unstable over the course of illness (Heslin et al., 2015), a transdiagnostic psychopathology approach was adopted during the recruitment phase. These cohorts were followed up approximately 5 years after their FEP, as part of the ‘Biological Phenotypes, Environment, Genes and Psychosis Outcome (GAP Follow up)’ study (Ethics ref 17/NI/0011).

Participants were eligible if they a) were aged between 18 and 65 years, b) presented with a clinical diagnosis of FEP, according to the ICD–10 criteria (Organization, 2004) (codes F20–F33), and c) resided within the defined catchment area. Participants were excluded if they: a) met the criteria for organic psychosis (F09), b) previously contacted mental health services for psychosis, and c) exhibited acute intoxication-related psychosis (ICD-10: F1X.5). Individuals without valid data on negative symptoms were further excluded from the analysis.

### Measures

The modified version of Medical Research Council (MRC) Sociodemographic Schedule (Mallett, 1997) was used to collect sociodemographic data and variables including employment, educational attainment, living and relationship status. We assessed six negative symptoms (restricted affect, diminished emotional range, poverty of speech, curbing of interests, diminished sense of purpose, and diminished social drive) of the participants at both time points using the Schedule for the Deficit Syndrome (SDS) on a scale of 4, where a score of 1 represents “not definitely absent” or “minimal presentation” on that symptom (Kirkpatrick et al., 1989).

Overall social function at each assessment point was assessed using the Global Assessment of Functioning (GAF) scale (Jones et al., 1995), ranging from 0 to 100, with higher scores indicating better function. The scores of positive and depressive symptoms were obtained from the transdiagnostic model of psychosis built on the Operational CRITeria (OPCRIT) system (McGuffin et al., 1991). Diagnoses were assigned according to the OPCRIT Research Diagnostic Criteria. General cognitive ability at baseline was assessed using the overall Intelligence Quotient (IQ) of the abbreviated Wechsler Adult Intelligence Scale(Blyler et al., 2000) (WAIS-III). Information on the duration of untreated psychosis (DUP) and hospitalization (e.g., days and times of hospitalization at follow up) were collected from the clinical records.

### Statistical Analysis

#### Sample descriptions

The prevalence of negative symptoms was assessed at baseline and follow-up. We applied growth mixture model (GMM), a statistical approach that estimates subgroups within a population based on their latent trajectories, to assign participants to four subgroups reflecting different baseline-to-follow-up profiles: a) *Subclinical*: no or minimum presence of negative symptoms at both time points, b) *Alleviating*: presence of negative symptoms at baseline but less so at follow-up, c) *Exacerbating*: no or minimum presence at baseline but worse at follow-up, and d) *Enduring*: presence of considerable level of negative symptoms across assessments. We compared baseline and outcome variables of the participants by these four trajectories using ANOVA and Post Hoc tests. The sample descriptions and analyses below (except for the factor analysis) were performed in IBM SPSS (version 28).

#### Factor Structure of negative symptoms

Confirmatory factor analysis (CFA) as implemented in the R-Package ‘lavaan’ (Rosseel, 2012) was employed to fit structural models, including: 1) a one-factor model reflects the global construct of negative symptoms; 2) a two-factor model with two uncorrelated negative symptoms factors (DE-AA); and 3) a correlated two-factor (DE-AA) model. To ensure equivalence in measures, configural invariance was examined by testing whether the same factor structure holds across baseline and follow-up. Metric invariance was then evaluated by constraining the factor loadings to be equivalent across time points.

The absolute fits of all the models were adjudged by the following statistics: a) Comparative Fit Index and Tucker-Lewis index (CFI & TLI; >0.95 indicating good model fit), Root Mean Square Error of Approximation and Standardized Root Mean square Residual (RMSEA & SRMR; <0.08 is considered reasonable fit) (Browne and Cudeck, 1992), and c) Normed Chi-squared (NC) (Bagozzi and Yi, 1988), which is less sensitive to the sample size (<5.00 is considered good model fit) (Schumacker and Lomax, 2004). Different models were compared using Akaike Information Criterion (AIC) and Bayesian Information Criterion (BIC), where lower values indicate better relative fits (Guthery et al., 2003).

The scores of individual negative symptoms were weighted by multiplying the factor loadings derived from the corresponding model before being incorporated into any latent factors of negative symptoms (see Supplementary Fig. 1). The total score representing the severity of global negative symptoms (SDS-Total) was calculated by summing the weighted scores of all six SDS items. The factor scores (SDS-DE and SDS-AA) were calculated by summing the weighted scores of the items loaded on each respective factor of negative symptoms.

#### Outcome Predictions

The factor scores of negative symptoms (SDS-DE and SDS-AA) at baseline were used as the primary predictors in all statistical models. In the analyses exploring the predictive value of the DE-AA structure in comparison with the unitary construct of negative symptoms, we replaced the factor scores with the score of total negative symptoms (SDS-Total).

Preliminary bivariate correlation and subsequent blockwise regression modelling were applied to examine the relationship between our predictors and the outcomes, including the generated four trajectories of negative symptoms, the GAF score, the length of hospitalization, and the quality-of-life outcomes. Covariates were selected based on theoretical relevance and prior evidence of their influence on negative symptoms, including sociodemographic (e.g., age, gender, ethnicity) and clinically relevant variables (e.g., DUP, IQ, positive and depressive symptom dimensions, and antipsychotic treatment in categories). Missing data were handled using multiple imputation to maintain statistical power.

We examined whether baseline scores of SDS-DE, SDS-AA, or SDS-Total were associated with 1) the aforementioned four trajectories of negative symptoms (subclinical, alleviating, exacerbating, and enduring) using logistic regression; 2) GAF scores at 5-year follow-up using linear regression; 3) the days and frequencies of hospitalization using Poisson regression, 4) dichotomous living conditions and relationship status using binary logistic regression, and 5) occupational status and educational attainment using ordinal regression.

## Results

### Sample Description

Of the 270 patients with FEP successfully followed up, 227 participants had valid measures of negative symptoms and were included in the analyses (see Supplementary Fig. 2 for exclusion process). Of these, N=170 (74.9%) completed negative symptom assessments at both baseline and follow-up (completers), while N=57 (25.1%) completed baseline assessments only (non-completers). No significant differences emerged between the two groups except for higher IQ and fewer days of hospitalization in the non-completer group (see Supplementary Table 1 for details). Regarding the prevalence of negative symptoms, 118 (52.0%) out of the 227 participants presented with at least one negative symptom at baseline. The prevalence decreased slightly to 47.6% after 5 years, with 81 out of 170 showing negative symptoms at follow-up (see Supplementary Table 2 for details).

The latent class analysis identified four distinct courses of change (Fig.1) as the optimal fit for negative symptoms (BIC: 3 classes -2240; 4 classes -2425; 5 classes -2075; see Supplementary Table 3 for details). The *enduring* course (28%, n=48) was characterized by considerable levels of negative symptoms at both baseline and 5-year follow-up. The *alleviating* trajectory (20%, n=34) represented those with moderate levels of negative symptoms at first onset which decreased to minimum levels at follow-up. The exacerbating trajectory (11%, n=19) displayed an increase of symptoms from minimum to moderate levels over time. The *subclinical* trajectory (41%, n=69) showed mild or minimum levels of negative symptoms throughout the 5-year follow-up.

**Fig. 1.**
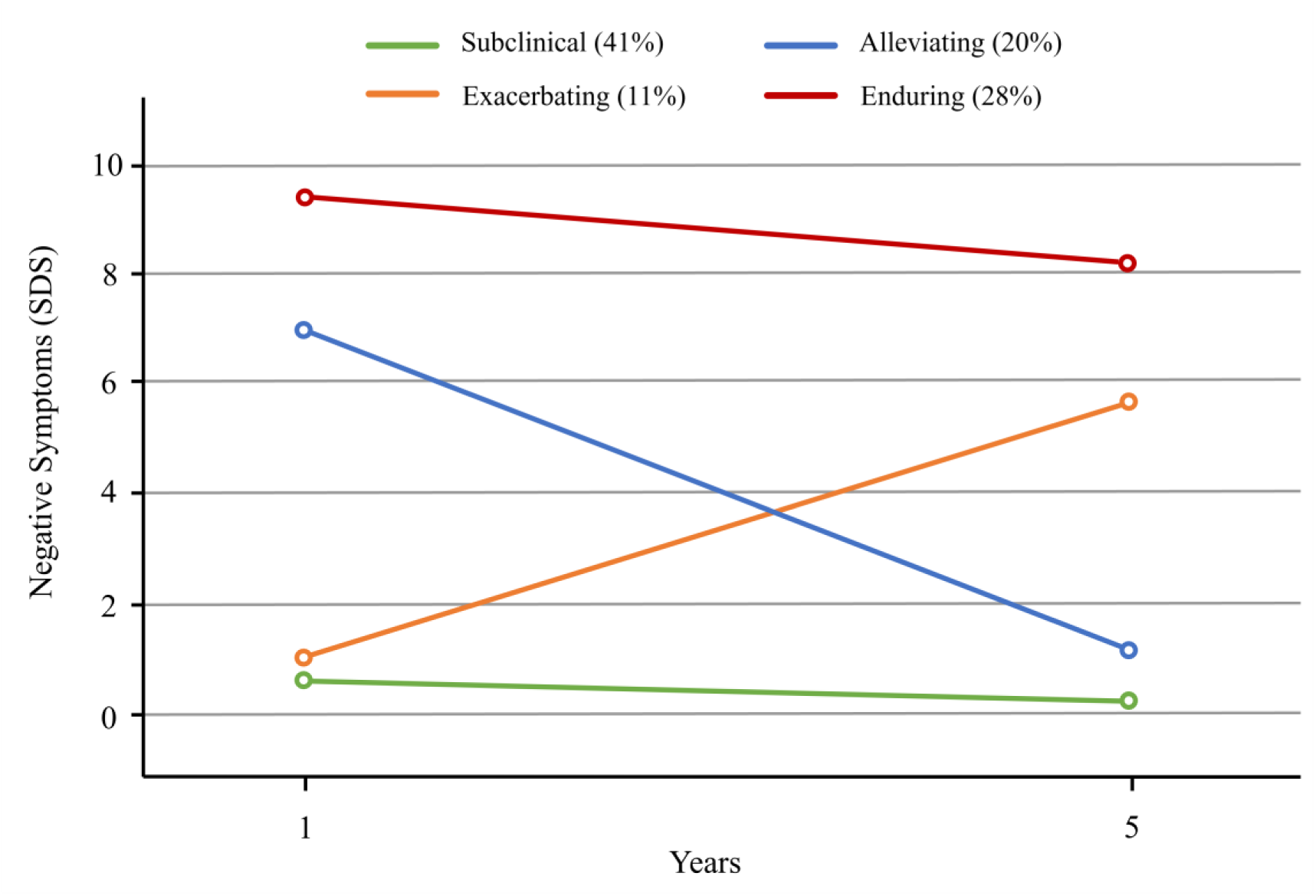
Four-course latent trajectories of negative symptoms, estimated by individuals’ scores performance on the Schedule of Deficit Syndrome (SDS) at both time points.

Table 1 summarizes the results of comparisons on baseline and follow-up characteristics across the four-trajectory groups of negative symptoms. Notably, more than two-thirds of participants in the enduring group were male. Compared with the subclinical group, participants in the enduring group exhibited significantly lower scores in baseline IQ (mean difference = 13.38 points) and poorer baseline functioning (GAF mean difference = 17.56 points). Post-hoc comparisons revealed that baseline AA—but not DE—significantly differentiated the enduring group from the alleviating group, with participants in the enduring group scoring on average 2.1 points higher in baseline AA (SE=0.57, 95%CI 0.93 to 3.88, p<0.001).

**Table 1.**
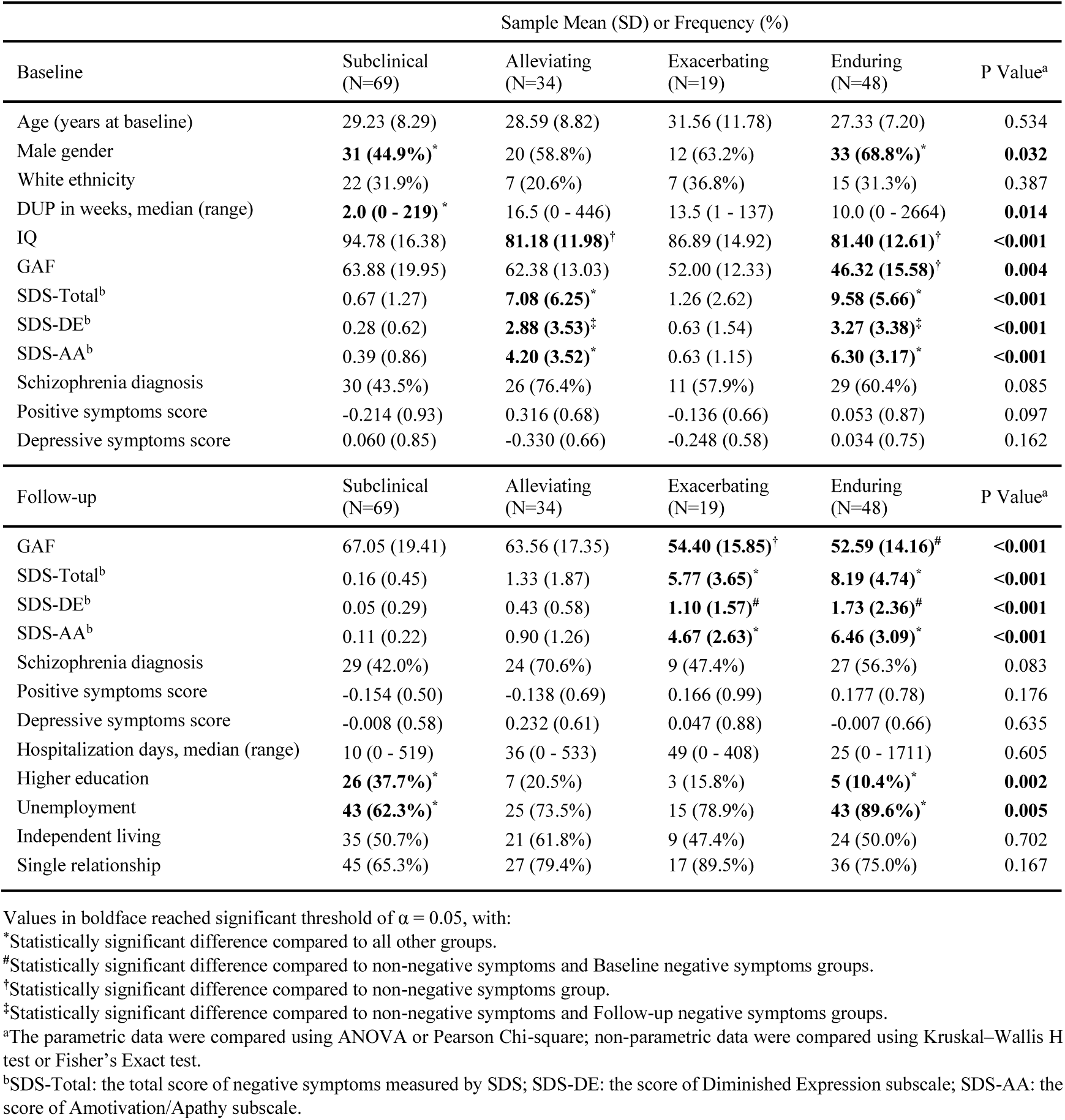
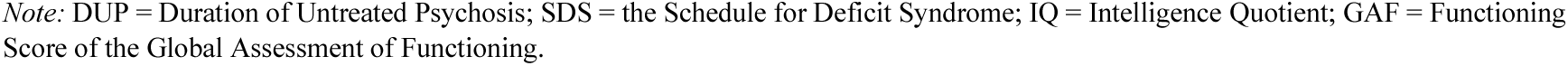
Comparisons of baseline and follow-up measurements across four trajectories of negative symptoms.

Additionally, individuals within the enduring trajectory group demonstrated clear disadvantages across multiple outcome measures, including poorer overall functioning, lower employment rates, and lower educational attainment compared to other trajectory groups. No cross-group differences were observed for the dimensional scores of positive and depressive symptoms at either baseline or follow-up assessments.

### Factorial Structure of negative symptoms

Compared to the one-factor and uncorrelated two-factor model, the correlated DE-AA model provided a better fit for negative symptom measures throughout assessments (Table 2; also see Supplementary Fig. 3 for all models being assessed). Both CFI and TLI values were higher than 0.95, indicating a good model fit, while NC and the SRMR values also met standard criteria for goodness-of-fit. RMSEA of the correlated two-factor model was within the acceptable range at baseline but slightly exceeded the threshold of 0.08 at follow-up. Nonetheless, tests of configural (CFI=0.995; TLI=0.994; RMSEA 0.016 [90%CI 0 to 0.054]) and metric invariance (Δχ² = 8.88, df = 8, p=0.35) indicated adequate longitudinal equivalence, demonstrating stability of the DE-AA structure across the assessment points.

**Table 2.** Fit statistics of negative symptom models from confirmatory factor analysis.

| Baseline Models<sup>a</sup> | Para | AIC | BIC | NC (df) | CFI | TLI | RMSEA | SRMR |
| --- | --- | --- | --- | --- | --- | --- | --- | --- |
| 1-Factor | 12 | 3502.06 | 3543.16 | 26.99 (9) | 0.831 | 0.718 | 0.318 | 0.091 |
| 2-Factor uncorrelated | 13 | 3463.85 | 3504.95 | 22.22 (9) | 0.862 | 0.770 | 0.287 | 0.400 |
| 2-Factor correlated | 13 | 3319.97 | 3364.49 | 3.99 (8) | 0.980 | 0.963 | 0.073 | 0.025 |
| Follow-up Models<sup>a</sup> | Para | AIC | BIC | NC (df) | CFI | TLI | RMSEA | SRMR |
| 1-Factor | 12 | 1952.70 | 1990.33 | 29.78 (9) | 0.742 | 0.570 | 0.387 | 0.167 |
| 2-Factor uncorrelated | 13 | 1804.50 | 1842.13 | 11.26 (9) | 0.909 | 0.848 | 0.230 | 0.309 |
| 2-Factor correlated | 13 | 1746.04 | 1786.81 | 3.70 (8) | 0.976 | 0.954 | 0.081 | 0.036 |

<sup>a</sup>The models included 1-Factor: the unidimensional model of negative symptoms; 2-Factor uncorrelated: the diminished expression (DE) and amotivation/apathy (AA) 2-factor model, with no correlation between the factors; 2-Factor correlated: the DE and AA 2-factor model, with correlation between the factors.
*Note:* Para = number of parameters; AIC = Akaike Information Criterion; BIC, Bayesian Information Criterion; NC = Normed Chi-squared; CFI, Confirmatory Fit Index; TLI, Tucker Lewis Index; RMSEA, Root Mean Square Error of Approximation; SRMR = Standardized Root Mean square Residual.

### Outcome Predictions

#### Persistence of Negative Symptoms

Regression analyses indicated that baseline AA, but not DE, significantly predicted the persistence of negative symptoms (Table 3). Higher AA scores at initial onset were associated with increased odds of exhibiting an enduring negative symptom trajectory over the 5-year follow-up. Male gender was also associated with slightly increased odds of being classified in the enduring trajectory group. After replacing the factor scores with the global score, baseline SDS-Total remained a significant predictor of enduring negative symptoms. However, the model including SDS-Total explained less variance compared to the model using the DE-AA dimensions as primary predictors.

**Table 3.**
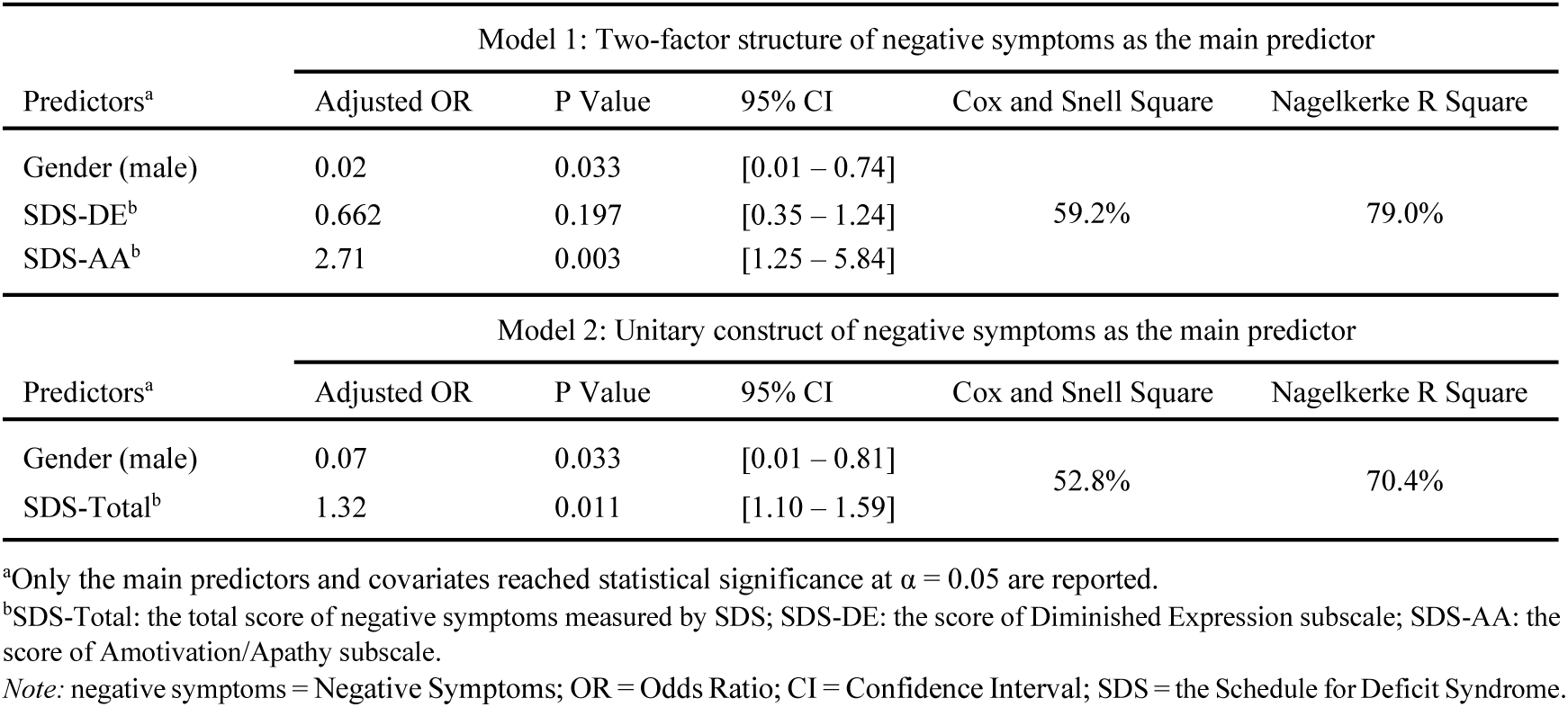
Effective predictors of the Two-factor and Unitary models in predicting the enduring course of negative symptoms.

| Model 1: Two-factor structure of negative symptoms as the main predictor |  |  |  |  |  |
| --- | --- | --- | --- | --- | --- |
| Predictors<sup>a</sup> | Adjusted OR | P Value | 95% CI | Cox and Snell Square | Nagelkerke R Square |
| Gender (male) | 0.02 | 0.033 | [0.01 – 0.74] |  |  |
| SDS-DE<sup>b</sup> | 0.662 | 0.197 | [0.35 – 1.24] | 59.2% | 79.0% |
| SDS-AA<sup>b</sup> | 2.71 | 0.003 | [1.25 – 5.84] |  |  |
| Model 2: Unitary construct of negative symptoms as the main predictor |  |  |  |  |  |
| Predictors<sup>a</sup> | Adjusted OR | P Value | 95% CI | Cox and Snell Square | Nagelkerke R Square |
| Gender (male) | 0.07 | 0.033 | [0.01 – 0.81] |  |  |
| SDS-Total<sup>b</sup> | 1.32 | 0.011 | [1.10 – 1.59] | 52.8% | 70.4% |

<sup>a</sup>Only the main predictors and covariates reached statistical significance at $\alpha = 0.05$ are reported.
<sup>b</sup>SDS-Total: the total score of negative symptoms measured by SDS; SDS-DE: the score of Diminished Expression subscale; SDS-AA: the score of Amotivation/Apathy subscale.
Note: negative symptoms = Negative Symptoms; OR = Odds Ratio; CI = Confidence Interval; SDS = the Schedule for Deficit Syndrome.

### Functional Outcome

Bivariate regression revealed significant associations of both baseline AA (B=-1.10, 95% CI -1.70 to -0.50, p<0.001) and total negative symptom scores (B=-0.53, 95% CI -0.90 to -0.16, p=0.005) with individuals’ GAF level at follow-up. Similar functional linkage was not detected with baseline DE (B=0.65, 95% CI -0.79 to 4.08, p=0.181). Multivariate models adjusting for baseline GAF further confirmed the directional association between baseline AA and functional outcome of the participants at follow-up (B=-1.76, 95% CI -3.40 to -0.11, p=0.037), where 46.1% of the total variance was explained by this adjusted model. However, after adjusting for baseline GAF, SDS-Total lost statistical significance in predicting the functional outcome of the individuals (B=-0.46, 95% CI -1.32 to 0.41, p=0.294), with baseline GAF retained as the only effective predictor (B=0.316, 95% CI 0.06 to 0.58, p=0.019).

### Hospitalization and Quality-of-Life Outcomes

None of the baseline measurements of negative symptoms were associated, at p=0.05 or less, with the length or frequency of hospitalization. Figure 2 compared the odds ratios (ORs) of the total and the factor scores of negative symptoms in predicting categorical quality-of-life outcomes. For occupational status, each 1-unit increase in baseline SDS-DE (OR=0.89, 95% CI 1.02 to 1.26, p=0.044), SDS-AA (OR=0.86, 95% CI 1.05 to 1.27, p=0.002), or SDS-Total (OR=0.92, 95% CI 1.03 to 1.15, p=0.005) significantly reduced the odds of employment at follow-up. Furthermore, higher baseline AA scores were associated with reduced odds of having an undergraduate or higher qualification (OR=0.93, 95% CI 1.04 to 1.16, p=0.019). None of the baseline measurements of negative symptoms effectively predicted participants’ living or relationship conditions after 5 years.

**Fig. 2.**
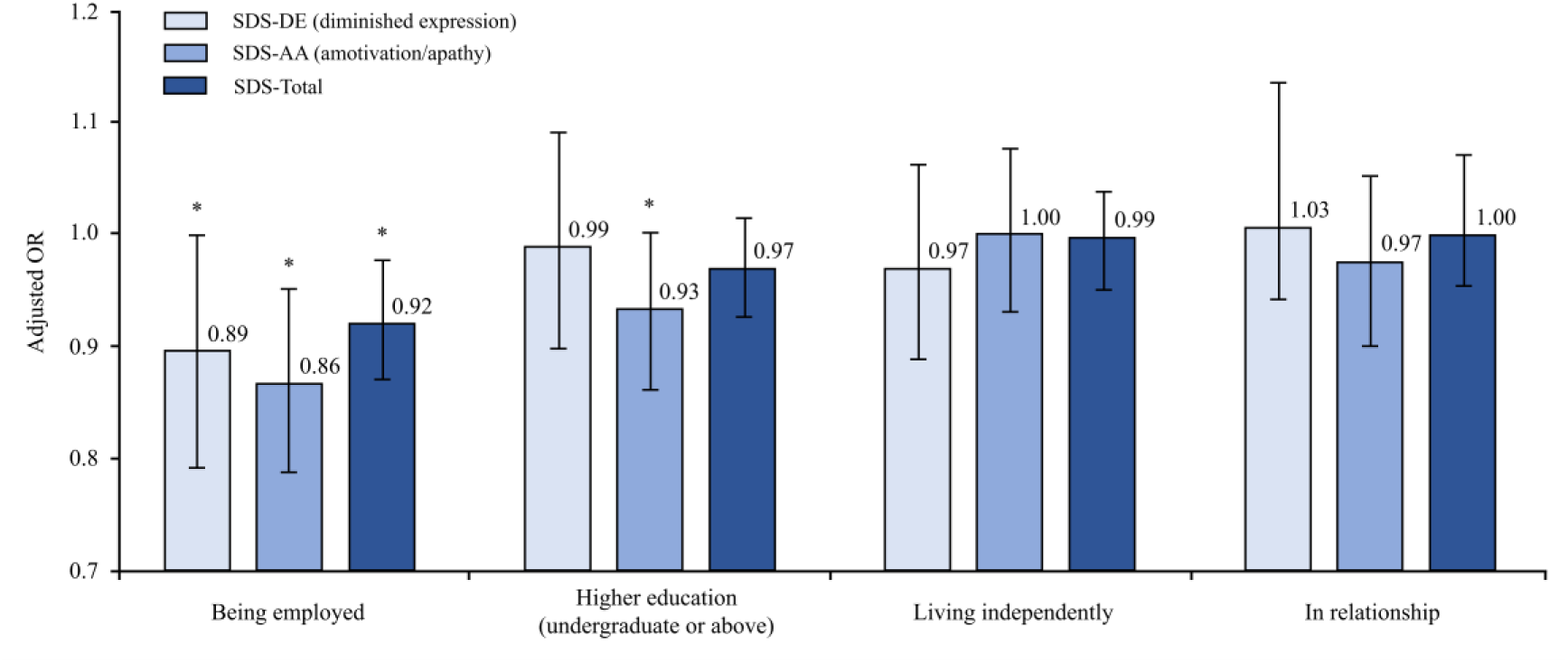
Predictive values of three negative symptom parameters on the ORs of four quality-of-life indicators (employment, educational attainment, living condition, and relationship status). Error bars represent 95% confidence intervals. *Asterisk indicates significant result at α = 0.05. SDS=the Schedule for Deficit Syndrome. OR=odds ratio.

## Discussion

### Main Findings

This is the first study to explore the longitudinal equivalence of the main factors of negative symptoms in FEP sample. First, we found that the correlated two-factor structure, composed of DE and AA, was evident at FEP and showed consistency across baseline and 5-year assessments. Second, baseline AA showed stronger predictive power than both DE and the unitary construct of negative symptoms in predicting whether symptoms persisted from baseline to 5-year follow-up. Third, with baseline functioning controlled, AA emerged as the only negative symptom dimension that significantly predicted participants’ functional outcomes at 5 years. Forth, all measures of negative symptoms showed an inverse relationship with individuals’ employment status at follow-up, while lower educational attainment was primarily associated with baseline AA.

### Comparison with Previous Research

Consistent with previous FEP studies, roughly half of our participants exhibited one or more negative symptoms at baseline. The presence of negative symptoms was associated with a higher proportion of males, longer DUP, and lower IQ, as extensively documented in previous literature (Ihler et al., 2021; Leeson et al., 2009; Ochoa et al., 2012). The slight decrease in the prevalence of negative symptoms at the 5-year follow-up contrasts the traditional view that negative symptoms accumulate with illness duration (Barrett, 1998). Additionally, the invariance of positive and depressive symptom scores across negative symptom trajectories supports the notion that negative symptoms represent a distinct psychopathological entity with respect to other psychotic symptoms (Foussias et al., 2014).

The factor structure of negative symptoms composed of DE and AA has been validated in cross-sectional studies of schizophrenia and schizoaffective cohorts (Kring et al., 2013; Strauss et al., 2012), and has also been supported by large-scale studies involving thousands of chronic patients across diverse geographical regions (Khan et al., 2017). Building on these findings, our study extends the dimensional approach to individuals experiencing their first episode of psychosis and further demonstrates the longitudinal equivalence of the DE-AA structure over a 5-year follow-up.

Previous studies have suggested that flat affect and alogia (components of DE) may not be the driving forces of the persistent nature of negative symptoms in FEP (Barch and Dowd, 2010). Our findings align with this perspective, highlighting that baseline AA emerged as the key predictor of enduring negative symptom trajectory. Moreover, individuals’ AA level at baseline outperformed DE in predicting their functional outcomes at follow-up. This is in accord with previous longitudinal and cross-sectional evidence linking motivational deficits closely to impaired functioning (Foussias et al., 2010). Such a prominent association between AA and functioning may already be established prior to the onset of psychotic episodes (Faerden et al., 2009).

Two possible explanations may account for the observed associations between AA, enduring negative symptoms, and poor functional outcomes. First, individuals with elevated AA levels may experience difficulties in initiating goal-directed activities such as help-seeking (Strauss and Gold, 2012). Similarly, motivational deficits could also lead to reduced treatment adherence (Cella et al., 2023). The lack of goal-directed behaviours and reduced compliance to treatment would then hinder symptomatic recovery and functional rehabilitation; Second, neurobiological dysfunctions involving the cortico-striatal circuitry might disrupt our reward-processing mechanisms. Although hedonic experience remains relatively intact (Burbridge and Barch, 2007; Dowd and Barch, 2010), individuals with schizophrenia may have difficulty in utilizing these reward information to modulate activities in the dorsolateral prefrontal cortex, resulting in a diminished motivational drive. Treatment with high doses of typical antipsychotics may further impede these reward-related processes (Juckel et al., 2006; Schlagenhauf et al., 2008).

Notably, after controlling for baseline GAF scores, participants’ AA levels still significantly predicted their long-term functional outcomes, demonstrating stronger predictive value than the total negative symptom score. A few studies have directly compared the predictive power between the subscales and broadly defined negative symptoms. For example, Harvey and colleagues (2017) found that the experiential factor of negative symptoms explained a greater variance in social impairment compared to either the expressive factor or the global negative symptom construct. This finding is consistent with our results, which demonstrated that baseline AA surpassed DE and the total negative symptom score in predicting GAF at the 5-year follow-up. It is therefore plausible that including DE—a negative symptom dimension less strongly related to functional outcomes (Green et al., 2012) —may dilute the predictive power of AA. Consequently, the global measure of negative symptoms assessed at FEP appears to have lost its value for long-term functional prognosis.

We found no significant association between negative symptoms and hospitalization among participants. Previous research suggests that more acute or prominent clinical issues (e.g., manic episode, substance abuse) tend to be the driving reason of hospital admission (Dollfus and Lyne, 2017; Patel et al., 2020), especially for individuals with FEP (Ajnakina et al., 2020). Additionally, social-environmental factors independent of symptomatic rehabilitation (e.g., self-caring capacity, social support, healthcare system differences) may also affect discharge process and lengthen the durations of hospitalization. We were unable to control for these contextual factors due to the practical challenges during data collection.

Regarding quality-of-life outcomes, higher scores on both AA and DE were slightly but significantly associated with lower odds of employment at follow-up. This is consistent with prior findings (Evensen et al., 2012) and further reinforcing the multifaceted nature of occupational success (Barch and Dowd, 2010). In contrast, educational attainment appears more dependent on one’s motivational implementation (Boekaerts et al., 2006) which primarily correlates with the presence of avolition, a core component of AA. However, we did not replicate previously reported associations between DE and individuals’ living and relationship conditions (Abplanalp et al., 2022). This discrepancy may be attributable to the relatively mild severity of DE observed in our first-episode sample, consistent with prior evidence suggesting that DE typically remains at lower levels among individuals with FEP (Peralta et al., 2014). Such mild symptom severity may have constrained our ability to identify associations previously observed in studies of chronic psychosis samples.

### Strengths and Limitations

First, our study validated the DE-AA structure of negative symptoms by extending its applicability to the early phase of psychosis. Second, to improve the accuracy of latent variable estimation, we weighted individual negative symptom items according to their factor loadings derived from the CFA results. Third, the longitudinal design allowed us to confirm the structural stability of the DE-AA dimensions over time and facilitated comparisons across distinct symptom trajectories. Additionally, our study directly compared the predictive utility of the two negative symptom dimensions versus their global construct, demonstrating the superior predictive value of AA in forecasting functional outcomes and symptom course trajectories.

This study has limitations. First, recent CFA and network analytic studies highlighted the possibility of underestimating the number of latent factors, especially when these factors are highly correlated (Strauss et al., 2019). Instead of the two-factor structure, researchers proposed better fits for the five-factor model and the hierarchical model (Ahmed et al., 2022). However, there is also evidence supporting the superior fit of the DE-AA structure even when assessed latent factors of negative symptom using second-generation scales (Li et al., 2022). Additionally, the number of latent factors identified in our study aligns with established recommendations for the item-to-factor ratio (Worthington and Whittaker, 2006), enhancing confidence in the robustness and reliability of the results.

Second, we cannot exclude the possibility that the strong linkage between AA and functional outcome was caused by the convergence of evaluation scopes rather than pathological causation per se. Like most traditional scales for negative symptoms, the SDS assesses AA primarily based on observable behavioural indicators. The assessment of observable behaviours, proposed in previous literature, can sometimes overlap with the assessments of functional capacity (Marder and Galderisi, 2017). Recent network analyses also reported that the motivational components of negative symptoms, such as avolition and anhedonia, may have blurred boundaries with functional domains in community detection (Abplanalp et al., 2022). Therefore, we encourage future studies using the second-generation scales of negative symptoms to distinguish AA from functional impairments from neurobiological and psychological perspectives.

Third, the absence of intermediate assessments between baseline and 5-year follow-up may limit our insights into the temporal progression of negative symptoms, particularly to any prominent change following the onset of the first episode. According to the “critical period” hypothesis (Birchwood et al., 1998), future research could consider adding additional assessment points within this timeframe to better understand how early interventions within the first year would affect the course development of negative symptoms.

### Implications

In line with current recommendations for evaluating negative symptoms (Galderisi et al., 2021; Kirkpatrick et al., 2006), our findings highlight the importance of assessing distinct symptom dimensions in both clinical and research contexts. The domain-specific approach underscores that AA may serve as a more clinically relevant marker than DE for prognostic prediction in FEP. Utilizing AA as a primary indicator, either alone or in combination with the unified construct of negative symptoms, could facilitate early identification of individuals at risk of persistent symptoms and poorer long-term outcome. The recognition of AA also sheds light on the development of targeted treatments. From an interventional perspective, motivation-oriented approaches—such as antipsychotics with minimal interference in dopaminergic reward-processing circuits (Juckel et al., 2006; Schlagenhauf et al., 2008) or motivation-focused psychotherapies—may improve rehabilitation outcomes in individuals with negative symptoms, and potentially support broader recovery across psychosis populations.

## Data Availability

All data produced in the present study are available upon reasonable request to the authors

## Supplementary Materials

**Supplementary Fig. 1.**
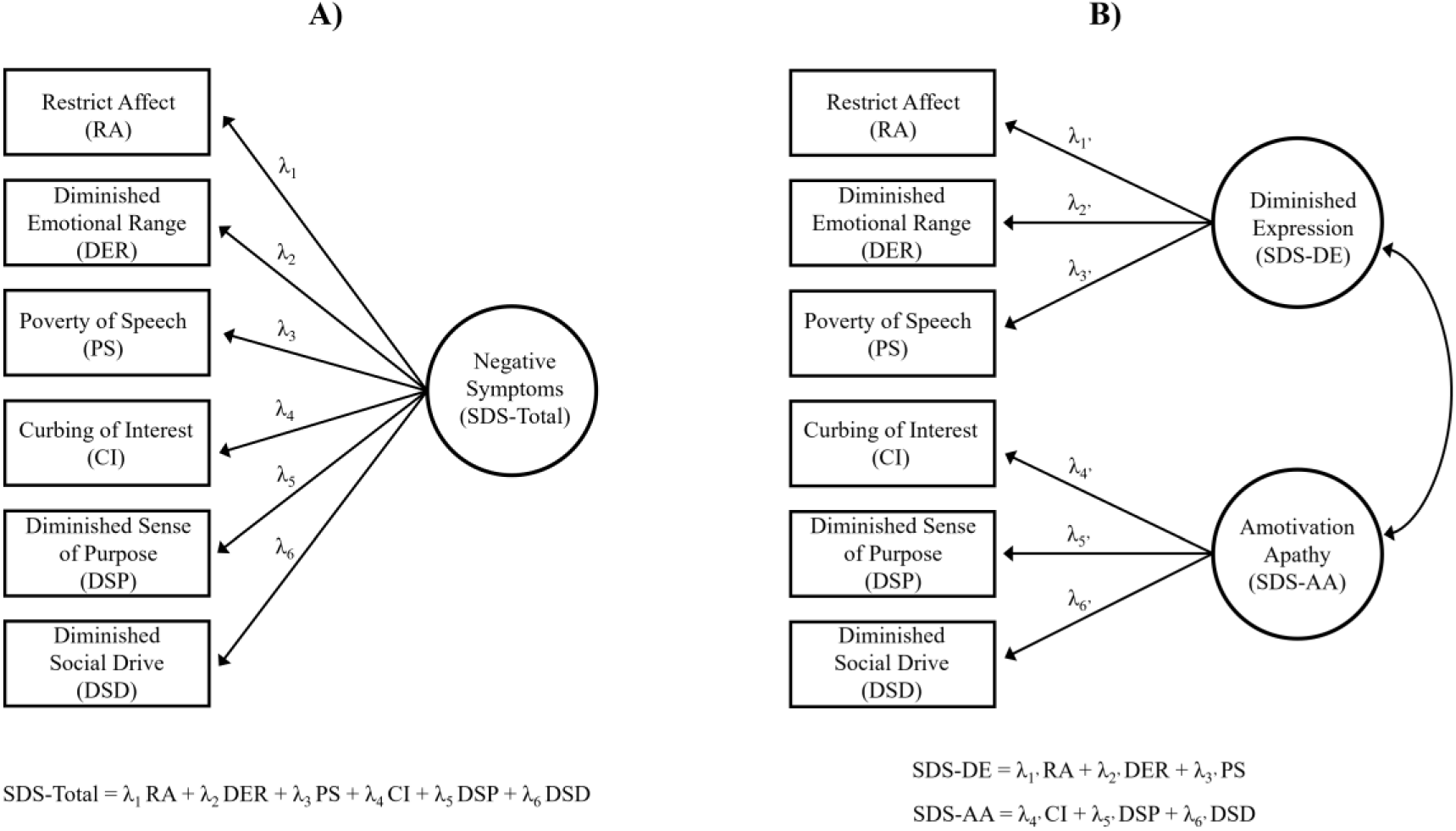
The demonstration of how A) the total score of negative symptoms (SDS-Total) was calculated based on the one-factor CFA model; B) the factor scores of negative symptoms (SDS-DE, SDS-AA) were calculated based on the two-factor correlated CFA models. The circles indicate the latent factors. The boxes indicate SDS items connected to their corresponding factors. The straight arrows indicate the loadings (λ) of individual items on their corresponding factors. The curve arrows indicate the covariances between the factors. The error terms of individual items are not shown in this figure for simplicity. *Note:* SDS = the Schedule for Deficit Syndrome; CFA = Confirmatory Factor Analysis

**Supplementary Fig. 2.**
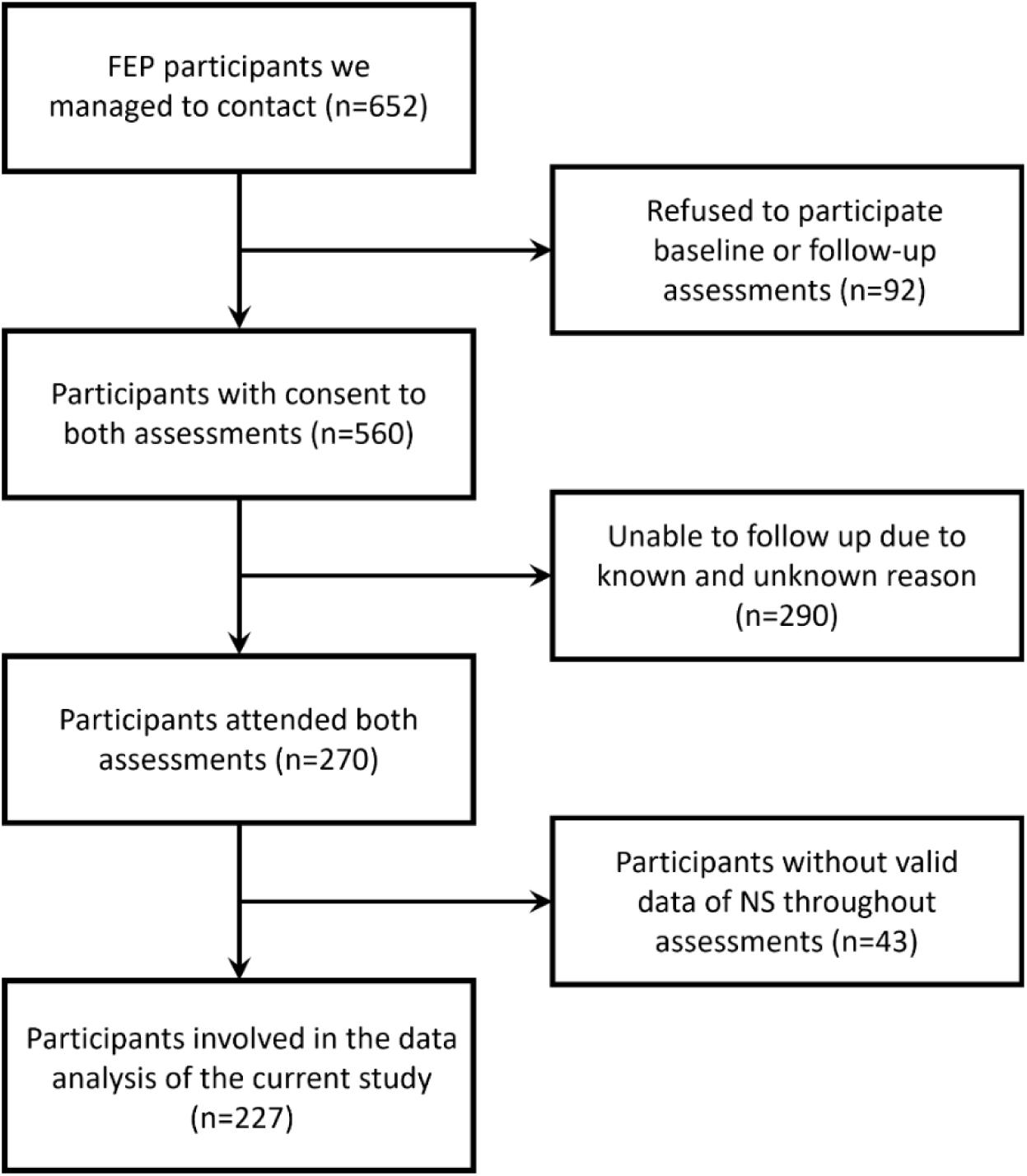
Exclusion flowchart for the participants involved in the data analysis of the current research. Out of the 652 FEP individuals we attempted to contact, 560 agreed to the participation of the study. Of the 290 participants who were not able to be followed up, 62 of them experienced geographic relocation such as moving or hospital transfer; 51 of them were not available due to special circumstances such as being significantly unwell/inpatient, imprisoned, homeless, or deceased; 177 of them were unable to attend/ to be contacted for unknown reasons. Individuals without valid data on negative symptoms were further excluded. *Note:* FEP = First Episode Psychosis.

**Supplementary Table 1.**

Within the 227 participants we identified with valid measures of negative symptoms for analysis, N=170 (74.9%) completed both baseline and follow-up measures of negative symptoms (completers), while N=57 (25.1%) only completed negative symptoms assessments at baseline (non-completers). No notable difference was observed between the two groups except for the higher IQ and lower days of hospitalization in the non-completer group

**Table 1.**
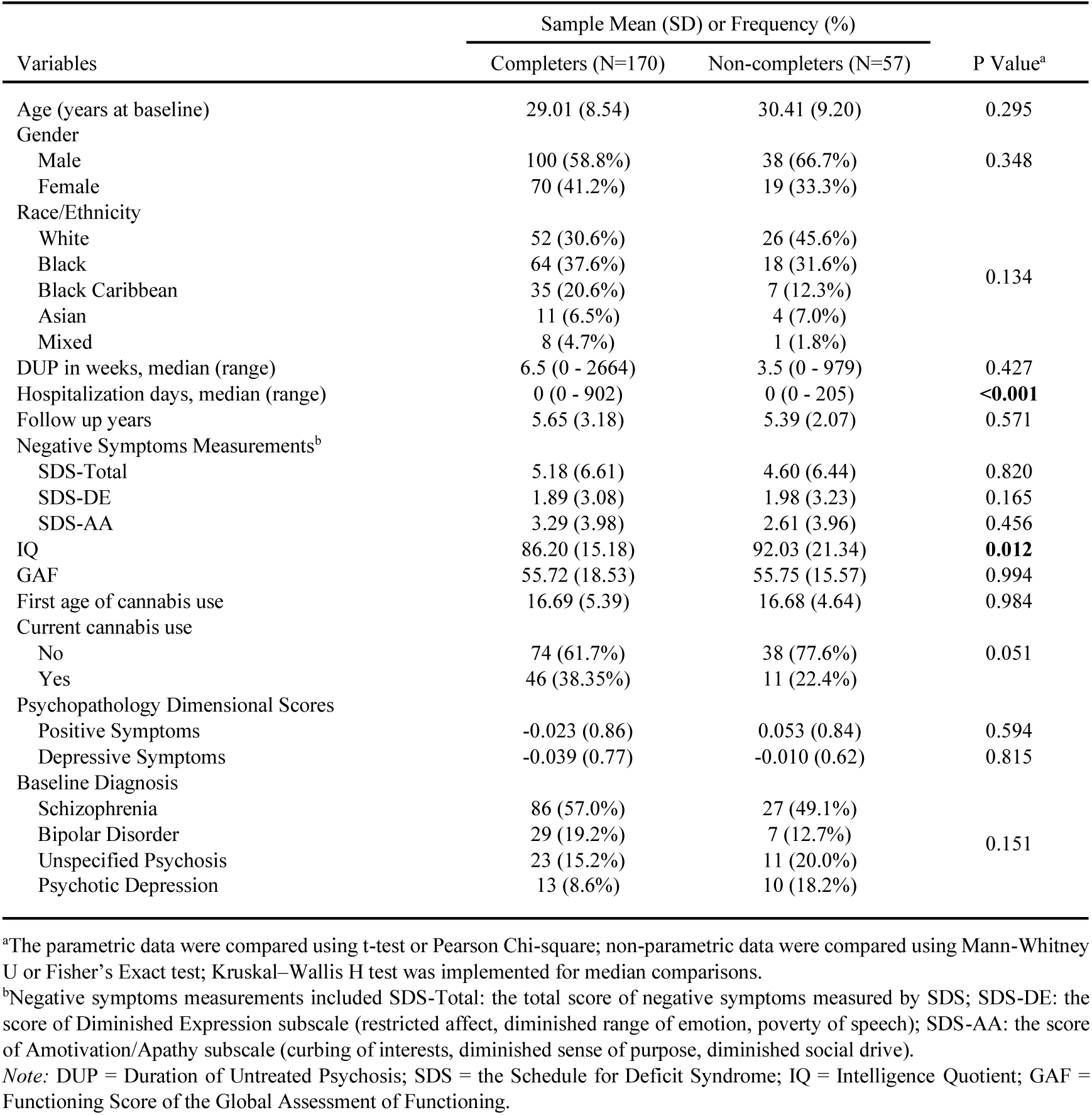
Demographic and clinical statistics of the study sample.

| Variables | Sample Mean (SD) or Frequency (%) |  | P Value<sup>a</sup> |
| --- | --- | --- | --- |
|  | Completers (N=170) | Non-completers (N=57) |  |
| Age (years at baseline) | 29.01 (8.54) | 30.41 (9.20) | 0.295 |
| Gender |  |  |  |
| Male | 100 (58.8%) | 38 (66.7%) | 0.348 |
| Female | 70 (41.2%) | 19 (33.3%) |  |
| Race/Ethnicity |  |  |  |
| White | 52 (30.6%) | 26 (45.6%) | 0.134 |
| Black | 64 (37.6%) | 18 (31.6%) |  |
| Black Caribbean | 35 (20.6%) | 7 (12.3%) |  |
| Asian | 11 (6.5%) | 4 (7.0%) |  |
| Mixed | 8 (4.7%) | 1 (1.8%) |  |
| DUP in weeks, median (range) | 6.5 (0 - 2664) | 3.5 (0 - 979) | 0.427 |
| Hospitalization days, median (range) | 0 (0 - 902) | 0 (0 - 205) | **<0.001** |
| Follow up years | 5.65 (3.18) | 5.39 (2.07) | 0.571 |
| Negative Symptoms Measurements<sup>b</sup> |  |  |  |
| SDS-Total | 5.18 (6.61) | 4.60 (6.44) | 0.820 |
| SDS-DE | 1.89 (3.08) | 1.98 (3.23) | 0.165 |
| SDS-AA | 3.29 (3.98) | 2.61 (3.96) | 0.456 |
| IQ | 86.20 (15.18) | 92.03 (21.34) | **0.012** |
| GAF | 55.72 (18.53) | 55.75 (15.57) | 0.994 |
| First age of cannabis use | 16.69 (5.39) | 16.68 (4.64) | 0.984 |
| Current cannabis use |  |  |  |
| No | 74 (61.7%) | 38 (77.6%) | 0.051 |
| Yes | 46 (38.35%) | 11 (22.4%) |  |
| Psychopathology Dimensional Scores |  |  |  |
| Positive Symptoms | -0.023 (0.86) | 0.053 (0.84) | 0.594 |
| Depressive Symptoms | -0.039 (0.77) | -0.010 (0.62) | 0.815 |
| Baseline Diagnosis |  |  |  |
| Schizophrenia | 86 (57.0%) | 27 (49.1%) | 0.151 |
| Bipolar Disorder | 29 (19.2%) | 7 (12.7%) |  |
| Unspecified Psychosis | 23 (15.2%) | 11 (20.0%) |  |
| Psychotic Depression | 13 (8.6%) | 10 (18.2%) |  |

<sup>a</sup>The parametric data were compared using t-test or Pearson Chi-square; non-parametric data were compared using Mann-Whitney U or Fisher's Exact test; Kruskal-Wallis H test was implemented for median comparisons.
<sup>b</sup>Negative symptoms measurements included SDS-Total: the total score of negative symptoms measured by SDS; SDS-DE: the score of Diminished Expression subscale (restricted affect, diminished range of emotion, poverty of speech); SDS-AA: the score of Amotivation/Apathy subscale (curbing of interests, diminished sense of purpose, diminished social drive).
*Note:* DUP = Duration of Untreated Psychosis; SDS = the Schedule for Deficit Syndrome; IQ = Intelligence Quotient; GAF = Functioning Score of the Global Assessment of Functioning.

**Supplementary Table 2.**

Cross-sectionally, participants were assigned to the group with or without at least minimal presentation of at least one negative symptom at each assessment point (NS and Non-NS groups). The socio-demographic, clinical, and outcome characteristics between the NS and Non-NS groups were summarised in Supplementary Table 2a (baseline) and Supplementary Table 2b (follow-up). At baseline assessment, the NS group had longer DUP, lower IQ, higher values on the dimensional score of positive symptoms, and higher rate of schizophrenia than the non-NS group. At follow-up, the NS group exhibited higher rate of unemployment, less educational attainments, and higher value of positive symptoms scores. Subjects with NS contained as significantly higher proportion of males and poorer GAF-F than the non-NS group throughout the assessments.

**Supplementary Table 2a.** The social-demographic and clinical characteristic comparisons between NS and Non-NS groups at baseline (N=227)

| Variables | Sample Mean (SD) or Frequency (%) |  | P Value<sup>a</sup> |
| --- | --- | --- | --- |
|  | NS group (N=118) | Non-NS group (N=109) |  |
| Age (years at baseline) | 28.64 (8.55) | 30.14 (8.85) | 0.147 |
| Gender |  |  |  |
| Male | 79 (66.9%) | 59 (54.1%) | 0.048 |
| Female | 39 (33.1%) | 50 (45.9%) |  |
| Race/Ethnicity |  |  |  |
| White | 37 (31.4%) | 41 (37.6%) | 0.659 |
| Black | 42 (35.6%) | 40 (36.7%) |  |
| Black Caribbean | 25 (21.2%) | 17 (15.6%) |  |
| Asian | 7 (5.9%) | 8 (7.3%) |  |
| Mixed | 6 (5.1%) | 3 (2.8%) |  |
| DUP in weeks, median (range) | 9 (0 - 2664) | 3 (0 - 630) | **0.019** |
| IQ | 89.56 (15.14) | 99.18 (16.44) | **0.004** |
| GAF-F | 50.69 (14.43) | 61.74 (19.07) | **0.024** |
| First age of cannabis use | 16.35 (4.57) | 17.06 (5.79) | 0.984 |
| Current cannabis use |  |  |  |
| No | 58 (65.2%) | 54 (67.5%) | 0.749 |
| Yes | 31 (34.8%) | 26 (32.5%) |  |
| Psychopathology Dimensional Scores |  |  |  |
| Positive Symptoms | 0.146 (0.84) | -0.168 (0.84) | **0.012** |
| Depressive Symptoms | -0.036 (0.72) | -0.025 (0.75) | 0.914 |
| Baseline Diagnosis |  |  |  |
| Schizophrenia | 71 (64.0%) | 42 (44.2%) | **0.011** |
| Bipolar Disorder | 12 (10.8%) | 24 (25.3%) |  |
| Unspecified Psychosis | 15 (13.5%) | 19 (20.0%) |  |
| Psychotic Depression | 13 (11.7%) | 10 (10.5%) |  |

Values in boldface reached significant threshold of $\alpha = 0.05$ .
<sup>a</sup>The parametric data were compared using t-test or Pearson Chi-square; non-parametric data were compared using Mann-Whitney U or Fisher's Exact test; Kruskal-Wallis H test was implemented for median comparisons.
Note: DUP = Duration of Untreated Psychosis; IQ = Intelligence Quotient; GAF-F = Functioning Score of the Global Assessment of Functioning.

**Supplementary Table 2b.**
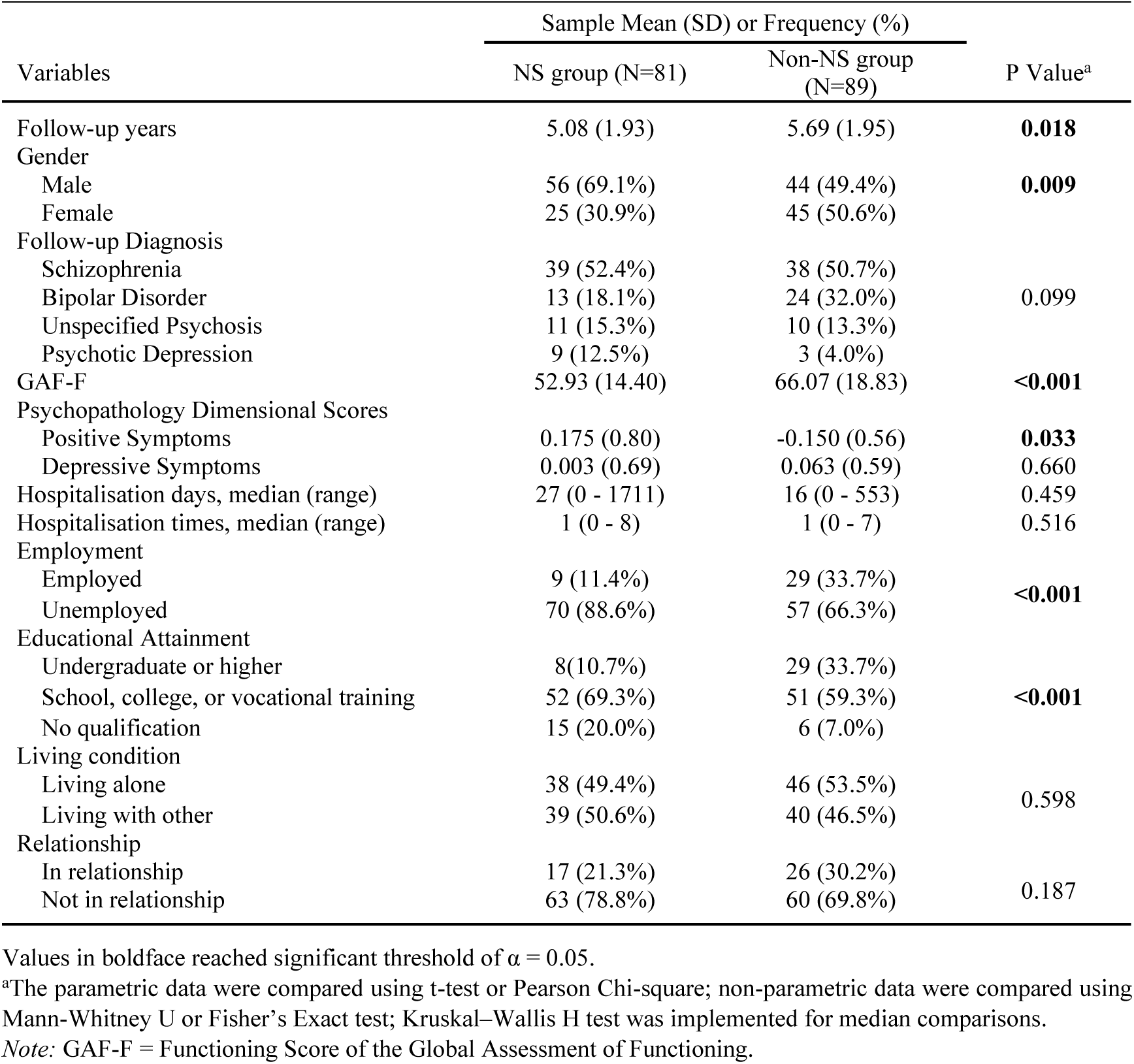
The social-demographic, clinical and outcome characteristic comparisons between NS and Non-NS groups at follow-up (N=170)

| Variables | Sample Mean (SD) or Frequency (%) |  | P Value<sup>a</sup> |
| --- | --- | --- | --- |
|  | NS group (N=81) | Non-NS group (N=89) |  |
| Follow-up years | 5.08 (1.93) | 5.69 (1.95) | **0.018** |
| Gender |  |  |  |
| Male | 56 (69.1%) | 44 (49.4%) | **0.009** |
| Female | 25 (30.9%) | 45 (50.6%) |  |
| Follow-up Diagnosis |  |  |  |
| Schizophrenia | 39 (52.4%) | 38 (50.7%) | 0.099 |
| Bipolar Disorder | 13 (18.1%) | 24 (32.0%) |  |
| Unspecified Psychosis | 11 (15.3%) | 10 (13.3%) |  |
| Psychotic Depression | 9 (12.5%) | 3 (4.0%) |  |
| GAF-F | 52.93 (14.40) | 66.07 (18.83) | **<0.001** |
| Psychopathology Dimensional Scores |  |  |  |
| Positive Symptoms | 0.175 (0.80) | -0.150 (0.56) | **0.033** |
| Depressive Symptoms | 0.003 (0.69) | 0.063 (0.59) | 0.660 |
| Hospitalisation days, median (range) | 27 (0 - 1711) | 16 (0 - 553) | 0.459 |
| Hospitalisation times, median (range) | 1 (0 - 8) | 1 (0 - 7) | 0.516 |
| Employment |  |  |  |
| Employed | 9 (11.4%) | 29 (33.7%) | **<0.001** |
| Unemployed | 70 (88.6%) | 57 (66.3%) |  |
| Educational Attainment |  |  |  |
| Undergraduate or higher | 8(10.7%) | 29 (33.7%) | **<0.001** |
| School, college, or vocational training | 52 (69.3%) | 51 (59.3%) |  |
| No qualification | 15 (20.0%) | 6 (7.0%) |  |
| Living condition |  |  |  |
| Living alone | 38 (49.4%) | 46 (53.5%) | 0.598 |
| Living with other | 39 (50.6%) | 40 (46.5%) |  |
| Relationship |  |  |  |
| In relationship | 17 (21.3%) | 26 (30.2%) | 0.187 |
| Not in relationship | 63 (78.8%) | 60 (69.8%) |  |

Values in boldface reached significant threshold of $\alpha = 0.05$ .
<sup>a</sup>The parametric data were compared using t-test or Pearson Chi-square; non-parametric data were compared using Mann-Whitney U or Fisher's Exact test; Kruskal-Wallis H test was implemented for median comparisons.
Note: GAF-F = Functioning Score of the Global Assessment of Functioning.

**Supplementary Table 3.**

The table below displays the classification distribution and model fit statistics for Models 1 through 5, as estimated using the growth mixture model. The three-class model (BIC: -2240) identified similar trajectories to the four-class solution but failed to differentiate an important subgroup. Specifically, it merged the exacerbating trajectory (which showed an increase in symptoms over time) with the subclinical trajectory, resulting in a broader group with heterogeneous symptom progression patterns. This oversimplification suggests that the three-class model lacked the granularity needed to capture the distinct worsening trend observed in a subset of individuals.

Conversely, the five-class model (BIC: -2075) introduced an additional class but did not substantially improve interpretability. Instead, it divided one of the existing trajectories, leading to an unstable classification where one class was rendered effectively redundant (0% assignment). Specifically, the new classification split the alleviating trajectory, creating an additional group with similar characteristics but no clear distinction in symptom progression. This overfitting issue, coupled with a higher BIC score, indicates that the five-class model imposed unnecessary complexity without offering meaningful insights.

**Supplementary Table 3.**
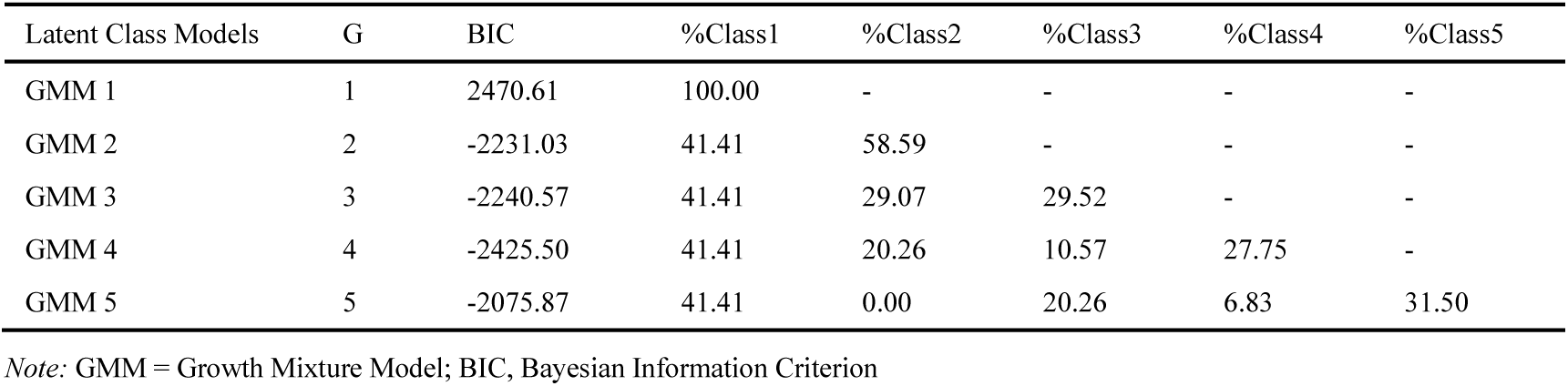
Classifications of latent trajectories of negative symptoms and fit statistics calculated by growth mixture model.

**Supplementary Fig. 3.**
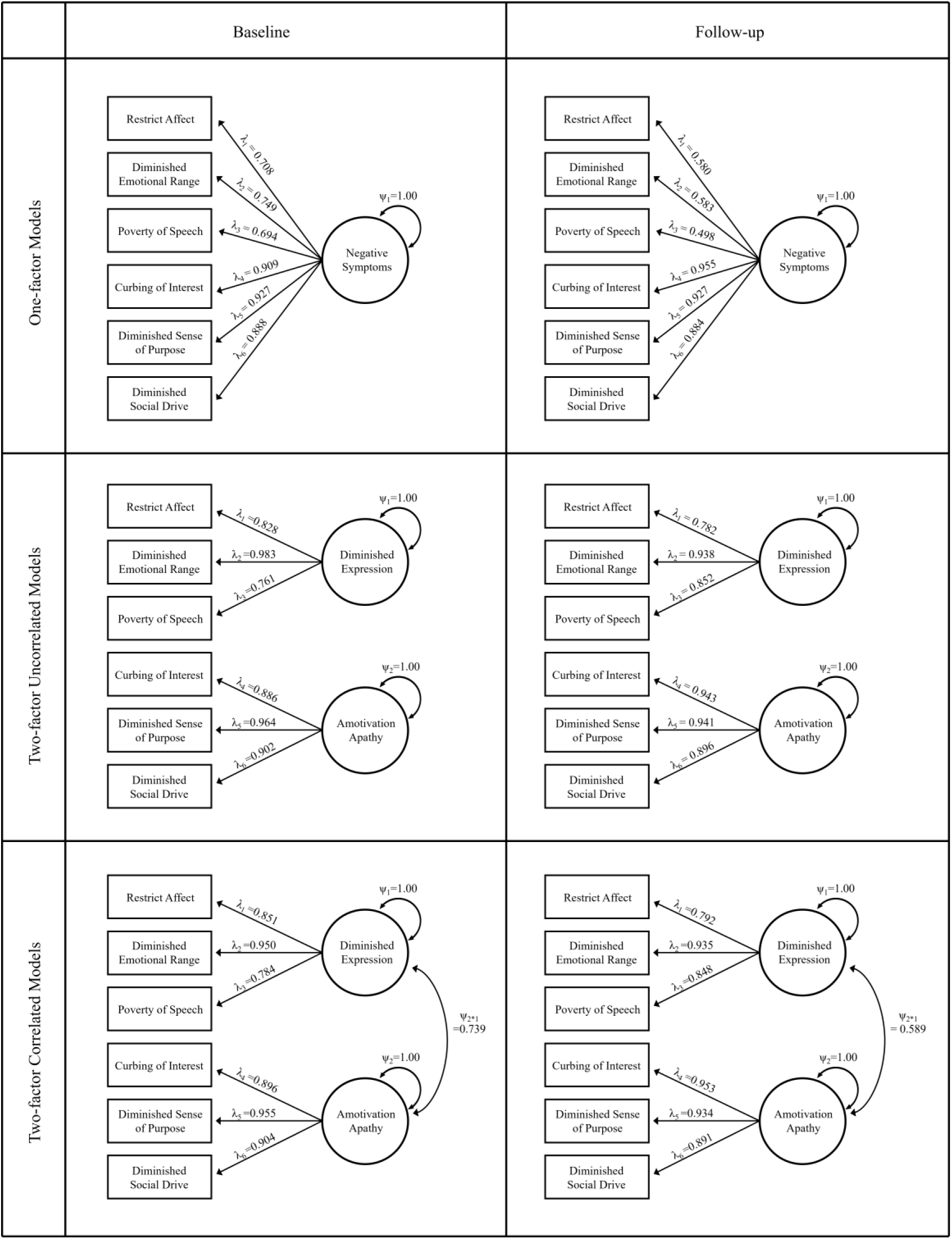
The one-factor (NS), two-factor uncorrelated (DE-AA), and two-factor correlated (DE-AA) models of negative symptoms at baseline and follow-up, with residual variances/covariances (ψ) and loadings (λ) assessed using confirmatory factor analysis. The circles indicate the latent factors. The boxes indicate SDS items connected to their corresponding factors. The straight arrows indicate the loadings of individual items on their corresponding factors. The curve arrows indicate the variances or covariances within or between the factors. The error terms of individual items are not shown in this figure for simplicity.

